# Sustainability of Evidence-Based Practice Improvement Programs in Abu Dhabi Ambulatory Healthcare Services for more than a decade and During the COVID-19 Pandemic

**DOI:** 10.1101/2025.03.11.25323728

**Authors:** Latifa Baynouna Alketbi, Nico Nagelkerke, Hanan Abdelbaki

## Abstract

The Abu Dhabi Ambulatory Healthcare Services (AHS) implemented the Chronic Disease Care (CDC) and Patient-Centered Medical Home (PCMH) programs. A retrospective observational descriptive design was used to analyze the sustainability of both programs. Linear regression showed that the key performance indicator (KPI) for the best-performing centers had significantly higher PCMH scores, with no effect on the financial revenue of the centers. Pearson correlation analysis indicated significant correlations between clinical and preventive KPI achievements and the 2022 PCMH and CDC scores. The AHS centers successfully implemented both programs sustainably. The study findings highlight areas for sustainability research that demonstrate the value of sustainable interventions.

**Contributions to the literature:**

- Deviations in achieving optimal healthcare outcomes are rooted in the lack of enough evidence-based interventions.
- Evidence-based interventions, like providing family medicine-based primary care, rank among the most thoroughly studied interventions.
- This study demonstrates the sustainability of the well-known, evidence-based intervention, NCQA PCMH standards.
- The established structure and processes for adapting the NCQA PCMH standards supported AHS centers during and after the COVID-19 pandemic, resulting in superior clinical and utilization outcomes in centers that implemented the standards more effectively.

## Background

Studies on healthcare services are neither designed nor funded for sufficiently long periods to examine adaptation, sustainability, or outcomes of healthcare changes or interventions over time (AHRQ, 2023). Thus, implementation science requires later-stage translation research questions for population impact. It is important to prioritize the current evidence gap in this area by focusing on the value of sustaining interventions over time, identifying sustainability correlations and strategies for sustaining evidence-based interventions. and advancing workforce capacity, research culture, and funding mechanisms (Proctor et al., 2015).

However, definitional issues regarding sustainability remain (Scheirer, Hartling, & Hagerman, 2008). Although sustainability can be broadly centered on whether an improvement program continues to exist (Chrysanthi Papoutsi, Sonja Marjanovic, 2024), a view of sustainability that is more akin to normalization has been adopted. In this view, “new ways of working and improved outcomes become the norm” without reverting to previous practices and maintaining better outcomes, which is critical in defining sustainability.

Many countries devote significant effort to the spread and scale-up of healthcare service improvements; however, a few of those that succeed locally are spread and sustained widely (Chrysanthi Papoutsi, Sonja Marjanovic, 2024). Another challenge in sustainability research is the ongoing change in the context and healthcare systems, increasing complexity of healthcare, resource constraints, growing diversity of populations, and the highly variable healthcare structure and process. Thus, healthcare leaders and managers rely on expertise to inform their decisions and, in many instances, for short-term gains mostly arising from problem situations (Macdonald, Bath, & Booth, 2008). Thus, developing and implementing cost-effective healthcare services without sustainability have contributed to the availability of evidence-based guidelines, standards, and policies that can improve healthcare but without sufficient evidence of their sustainability. This dearth in sustainability research prompts epistemological and methodological questions (Greenhalgh, Macfarlane, Barton-Sweeney, & Woodard, 2012; Scheirer, 2005)

This study describes an experience of sustaining healthcare programs in a new context and country. Abu Dhabi Ambulatory Healthcare Services (AHS) sustained the implementation of two best evidence-based standards for the provision of primary care for over a decade. They are the Chronic Disease Care (CDC) program based on the chronic care model (CCM) by Wagner, implemented in 2004, and the Patient-Centered Medical Home (PCMH) program by the National Commission for Quality Assurance (NCQA), implemented in 2013 (NCQA,; Wagner et al., 2001). The goals of both programs align and overlap in terms of time and shared components. This provides additional learning potential for practice improvement through multiple solutions with changing contexts and challenges. Both programs have been reported in previous publication (Baynouna Al Ketbi et al., 2018; Baynouna et al., 2010), and their impact has been reviewed (Paulo, Loney, & Lapão, 2017).

The CCM, a chronic disease management program, was conceptualized and implemented in primary care in 1998. The most known framework is the CCM by Wagner, who summarized the best research evidence in the CCM to guide quality improvement. The effectiveness of this model was evident in processes, health services, quality of life (QoL), health outcomes, satisfaction, and costs, in addition to an observed decrease in coronary heart disease mortality (Wagner et al., 2001) (Bodenheimer, Wagner, & Grumbach, 2002). In a systematic review of 77 papers on the effectiveness of the CCM, all but two reported improvements in healthcare practice or health outcomes for people with chronic diseases. The systematic review revealed hidden determinants (Davy et al., 2015), including leadership commitment, awareness of end-users and reflective healthcare practice, and leaders’ support of the implementation and sustainability of interventions, which were just as important as the CCM elements.

The NCQA program (NCQA,) has six well-established best-evidence standards. It covers service provision in primary care based on family medicine principles. The standards are focused on team-based care, knowing and managing populations, access and continuity, care management, care coordination, and performance measurement. Its effectiveness in delivering high-value care has been reported (Grove et al., 2020; Swietek et al., 2018; Rosland et al., 2018; Baynouna Al Ketbi et al., 2018; Burton, Zuckerman, Haber, & Keyes, 2020; John, Jani, Peters, Agho, & Tannous, 2020; NCQA, 2019; Quigley, Slaughter, Qureshi, Elliott, & Hays, 2021; van den Berk-Clark et al., 2018) (Shi et al., 2017) (Nelson et al., 2014).

This study examines the long-term sustainability of the CCM-based CDC and NCQA PCMH programs. Additionally, it outlines the impact of the COVID-19 pandemic as a disaster in healthcare on both programs. The successful effects of these programs on patient outcomes developed in the US and their transferability can offer insight for generalizability.

## Method

This study employed a retrospective observational descriptive design.

### Setting

Abu Dhabi citizens register with AHS in primary healthcare centers. Comprehensive healthcare services in family medicine include urgent, chronic, and preventive services, as well as specialty services, such as pediatrics, obstetrics, and gynecology. The centers are equipped with in-center imaging, x-ray and ultrasound, pharmacy, laboratory, and dental facilities. The payment system is an open-access fee for services covered by the government for United Arab Emirates (UAE) nationals. Non-UAE nationals or their employers cover health insurance plans. Family physicians, general practitioners, and specialists provide care. The population and healthcare system of Abu Dhabi have been described (Koornneef, Robben, & Blair, 2017) (Paulo et al., 2017) ((DOH), 2018).

### Governance structure

In Abu Dhabi, healthcare facilities are governed by a regulatory body, the Department of Health (DOH), and healthcare service centers and hospitals are the operators. The AHS network of primary healthcare centers is linked to six government hospitals for secondary care. For patient care, AHS and all hospitals are linked with one Electronic Medical Record (EMR) introduced in 2009. The DOH publishes policies and screening and management guidelines. It oversees the implementation of national health programs and any cost coverage from the government. In addition, AHS implements certain programs as practice improvement initiatives (Appendix 1). PCMH and CDC programs have been overseen by the Academic Department through the Practice Improvement Committee since 2004. The Committee has been reporting to the PCMH steering committee since 2013.

### Communication and training

Communication with the healthcare centers occurs through facilitators in regular practice improvement meetings where key AHS staff are invited. At least once annually, a collaborative meeting is conducted to share the experiences and success of the center teams in CDC and PCMH implementation. An annual PCMH conference is conducted with experts in PCMH from the United States. Organizational training is provided for the center teams through the Continuous Professional Department in Academic Affairs.

### Chronic Disease Care Program

The CDC program was established in 2004. It boasted a system change in the centers in terms of structured daily clinics for patients with chronic diseases and a fixed schedule of the trained healthcare team led by a physician. In addition, EMR was continuously optimized to meet the needs of patients. Biannually, a comprehensive audit is conducted to benchmark the centers against each other according to the standards implemented.

### Patient-Centered Medical Home Program

Based on NCQA PCMH standards and the self-assessment that identified AHS gaps toward meeting the standards, a road map with multiple initiatives was developed. For each AHS department, medical, nursing, patient experience, pharmacy, dental, school health, health informatics, registration, allied health, and quality tasks were communicated and followed. The progress and approval of changes were tracked by a steering committee in the medical office for PCMH issues. Progress was based on the six NCQA standards and their updates and extended to areas targeted because of the local settings and unique opportunities for improvements. An example of modification is the requirement of a Primary Care Dentist (PCD) and the development of a population and panel management report from the EMR. Similarly, each school had a primary school nurse, an empanelment was performed, and reports were extracted for the panel management of school nurse students. Elements in the PCMH are presented in Appendix 2.

### Program facilitation

The project lead and two full-time facilitators (practice improvement and care coordination facilitators) were responsible for facilitating and monitoring the program. The practice improvement facilitator is responsible for the agenda for implementation. She conducts site visits to all centers at least four times a year and communications through calls, virtual meetings, and emails. The central practice improvement committee approves an agenda, which the practice improvement facilitator delivers to the centers during a prearranged site visit. The visit can be geared toward training and informing the center teams of new changes or projects or auditing to ensure adherence to standards. The main agenda of the facilitation visits is a new addition to the program. Many ideas come from the center teams, and if suitable, are disseminated after approval by the practice improvement committee.

### Care coordination

The care coordination facilitator is responsible for care coordination elements in the NCQA PCMH standards, including empanelment, population health programs, core coordination care management, and PCMH metrics. She works on the EMR by coordinating its building, validation, extraction, and dissemination. Each center must have one care coordinator. She trains care coordinators and supports their ongoing queries. She audits their work and implements care coordination tasks for patients through the following changes and implemented strategies:

1. Maintaining the patient panel of the primary care physician (PCP) and continuous cleanup of such panels.
2. Population health lists are provided to the centers monthly to coordinate care and management. The programs implemented are Chronic Kidney Diseases, Ischemic Heart Diseases, Stroke, Osteoporosis, Dyslipidemia, Undiagnosed Hypertension, and Prediabetes.
3. Tracking the use of EMR reports and safety initiatives, such as investigations, referrals, and medication safety KPIs.
4. Program-specific metrics are prepared by the care coordination facilitator and published to all centers monthly, in addition to the KPI of the institution.

### Use of Electronic Medical Records and program metrics

Both programs were facilitated by different supporting factors. First, the unified integrated Health Information System with the accumulative patient information data is a tool used to build and generate reports to identify different populations. It is the main method for identifying and tagging populations. Second, patient stratification based on published strategies (Reddy et al., 2017) was implemented over the years based on either disease, population encounter, or global risk assessment. The disease stratification was based on diabetes, hypertension, asthma, prediabetes, ischemic heart disease, stroke, or chronic kidney disease (CKD). Another supporting factor was the national government programs with patients registering for preventive services, such as premarital, well-childcare, school screening, cardiovascular screening programs, or cancer screening, which increased patient engagement with the centers.

A global risk assessment was performed by PCP clinical institution based on a modified American Academy of Family Physicians (AAFP) risk stratification (Stratification,). The stratified groups of patients were recalled and booked for appointments with the PCP for proactive care, disease management, and the identification of uncontrolled high-risk patients in their panel. EMR reports were used to monitor implementation, and feedback to the AHS centers and PCP is sent monthly.

### Data collection

This study used routinely gathered KPIs for the CDC and PCMH. Both programs assessed and tracked many outcomes. To simplify the analysis, the results were presented as measures of adherence to both programs (the accumulative annual score of the PCMH assessment and the CDC annual assessment score). Key Performance Measures related to care outcomes were grouped into preventive, clinical, utilization, and processes.

Most of the preventive and clinical KPIs were published monthly by the quality department. They included metrics on diabetes mellitus, cancer screening preventive programs, hypertension, depression and anxiety screening, and asthma. The central implementation committee developed additional clinical KPIs to monitor the newly implemented population health and disease Management programs were also published monthly. These included CKD, heart and stroke, smoking cessation, prediabetes, dyslipidemia, complex patients, and obesity. Other KPI processes related to program implementation were medication reconciliation, referral feedback, investigation tracking, message center utilization, use of point-of-care decision support, risk stratification rate, departmental summary generation, primary dentist assignment, and PCP empanelment. Utilization KPIs included calling patients who were admitted or visited the emergency room, panel management by the PCP, and panel size adjustment.

Two major audits were conducted biannually to assess compliance with the CDC and PCMH programs in May–June (mid-year) and December–January (end-year). The PCMH NCQA self-assessment audit is a score of the accumulation of points gained by the center for meeting elements in the six PCMH standards. For example, the Care Management and Support (CM) standard has two competencies, A and B weighing 10 points of the total score (four core, all must be met, and six electives, few can be not met). Competency A weighed six points for three elements to be met, two points for two core elements, and two for one elective element. Appendix 2-A lists all points distribution; all points were added together with the AHS centers required to achieve 97 points. Although the total points required by the NCQA tool was 123, the AHS program required 97 as there were elements in the program which still in progress of development by the Practice Improvement Committee or it was not yet available in the AHS. Examples are two-way electronic communication between patients and healthcare providers, systematically obtaining prescription claims data to assess and address medication adherence, and publicly reporting clinician performance. The audit is performed by the program facilitators, and evidence is required for each element, which renders the assessment more objective. Sustainability was assessed over the years with the persistence of adherence to the elements of the standards and evidence provided to earn score points. The present NCQA self-assessment was based on the 2017 NCQA PCMH standards, although an older version based on the 2011 and 2014 NCQA standards was previously used. The CDC program employed an internally built audit tool (Appendix 2-B) based on earning points for adherence to the elements that were developed over the years by the Practice Improvement Committee and as well evidence is required. The results of both audits are distributed to all centers for action plans and benchmarking.

The data and evidence were EMR extracts, audits from EMR reviews, or field verification by the auditor. Very few elements were manually audited, e.g., care plan documentation. Centers receive the results and can provide feedback on the data or KPI validity as they have the raw data, and they can identify patients with gaps and verify the accuracy of such gaps.

### Awareness program

The PCMH program was identified as “Baytona Altebi,” meaning “Our Medical Home.” A continuous awareness program began since the launch of the PCMH program in 2013 for AHS employees and patients under this name. The launch of the employee program was through the first PCMH conference attended by senior leadership and officially announced as a government-supported initiative. Speakers with depth expertise in PCMH were invited. The community awareness is continuous, accompanied by logos, and placed on the buildings of all centers and educational material. Educational materials were developed, and the patients received PCP letters to introduce them to the PCP concept, accountability, and continuity. The CDC program was identified by this abbreviation, and within the EMR system, care was structured under this name.

### Ethics approval and consent to participate

The study was approved by the DOH-Abu Dhabi Institutional Review Board (IRB). All the methods were conducted following relevant guidelines and regulations. Informed consent was waived by the DOH-Abu Dhabi IRB, as the study was designed for retrospective data gathered as part of a patient care and organization quality program and anonymized during analysis.

### Data analysis

KPI and audit data were summarized, graphed, analyzed, and reported using Microsoft Excel® (Microsoft, Redmond, WA) and SPSS Version 29. Descriptive statistics, frequency, and the percentage of adherence to PCMH and CDC standards were calculated. Univariate and multivariate linear regressions were employed to assess the association between the performance outcomes related to the KPI and the PCMH and CDC scores. The multivariable model was adjusted for variables such as financial performance, city, and being a rural center. Pearson correlation analysis (PCA) was employed to assess the correlation of PCMH and CDC scores with other important variables, such as the centers’ financial revenue, laboratory imaging, and pharmacy orders. Statistical significance was set at p < 0.05.

## Results

### Population

In AHS centers, females were more frequent attendees, 60.9% compared to 39.1% among males. Children less than 18 years constitute 42.6% compared to 46.5 in the age group between 18 years and 59 years. Patients aged 60 years, or more are 10.9% of the attendee. Mainly UAE nationals were the main visitors to the centers. The average centers’ financial revenue in 2021 and 2022 was mainly from consultation visits (79.9%), preventive visits (15.8%), and teleconsultations (4.2%). It varied by the sizes of the centers, the 26 centers included seven rural centers, which had a considerably low number of patients.

### Program adherence

Most of the 26 centers implemented both programs well, with excellent sustainability and progress over time. Table 1 shows a drop in the PCMH scores and KPIs during the COVID-19 pandemic. Furthermore, Table 1 shows the CDC program adherence score and the score of the NCQA self-assessment audit in 2018, 2021, and 2022.

**Table 1.** Description of the assessment variables used in studying the value and sustainability of the NCQA PCMH and the chronic disease care standards in the 26 centers.

|  | No. of centers | Minimum | Maximum | Mean | SD |
| --- | --- | --- | --- | --- | --- |
| PCMH Audit |  |  |  |  |  |
| PCMH NCQA. End 2018 | 26.0 | 54.0 | 83.0 | 75.3 | 6.2 |
| PCMH NCQA. End 2021 | 26.0 | 66.0 | 81.0 | 72.8 | 4.2 |
| PCMH NCQA. End 2022 | 26.0 | 77.0 | 88.0 | 83.5 | 3.5 |
| Chronic Diseases Care Audit |  |  |  |  |  |
| CDC audit. End 2022 | 26.0 | 30.0 | 90.3 | 73.3 | 15.6 |
| Clinical Key Performance Indicators aggregate percentage |  |  |  |  |  |
| Clinical KPI. End 2021 | 26.0 | 50.5 | 73.2 | 64.9 | 5.8 |
| Clinical KPI. End 2022 | 26.0 | 51.0 | 77.0 | 63.9 | 7.0 |
| Clinical KPI. Mid 2023 | 26.0 | 57.3 | 84.3 | 72.9 | 6.4 |
| Preventive Key Performance Indicators aggregate percentage |  |  |  |  |  |
| Preventive KPI. End 2022 | 26.0 | 33.0 | 58.0 | 47.8 | 7.0 |

In 2021, the best-performing center regarding PCMH implementation based on the NCQA PCMH score audit was achieving 83.5% of the best possible score (81 points out of a possible 97), while the worst-performing was 68% (66 out of 97). In 2022, with more efforts toward recovery from the pandemic, the best performing was 90.7% (88 out of 97), and the worst was 79.4% (77 out of 97). A clear improvement was observed in 2022 following very difficult years of AHS services directed toward COVID-19 prevention and mitigation. Compared to the first NCQA assessment done in AHS in 2018 based on the 2017 standards, the best that could be achieved in AHS was 83 points (92.2%), and the worst was 54 (58.7%). In 2022, adherence to structured care in the management of chronic diseases was 73.3% on average, with top-performing centers achieving 90%. Achievement of the best targets in clinical and preventive KPI for the year 2022 was 77% and 58%, respectively. The clinical KPI showed persistent improvement from 2021, from 73.2% in 2021 to 77% in 2022 to 84.3% in 2023. (Table 1).

### PCMH and CDC scores in relation to KPI and utilization

Figure 1 shows the NCQA PCMH assessment scores over 2018, 2021, and 2022 and the implementation of chronic disease management program standards related to three clinical KPI achievement categories: low (<60), moderate (60–70), and high performers (>70).

**Figure 1a.**
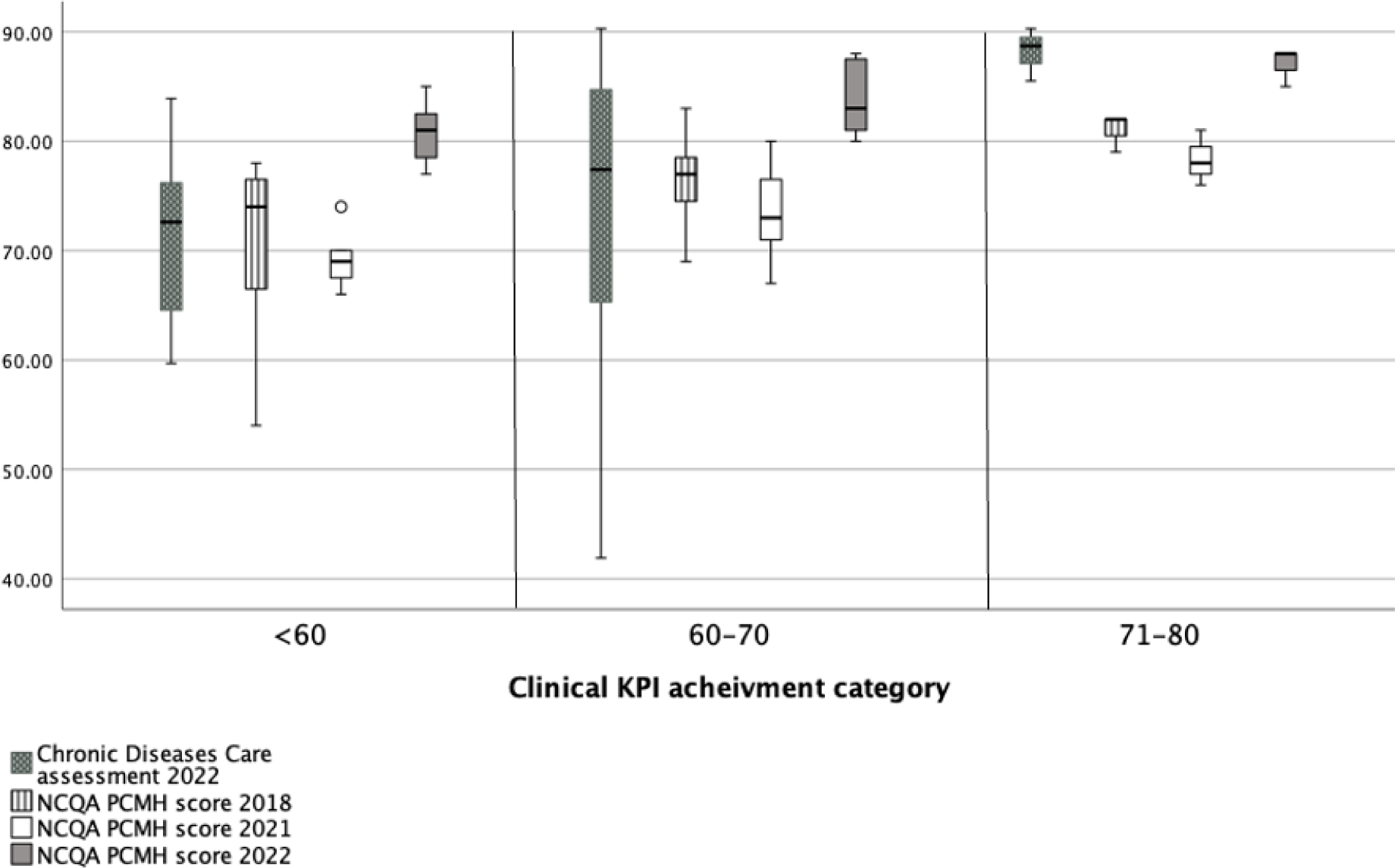
NCQA PCMH assessments’ scores over the years 2018, 2021, and 2022 and the chronic disease management program standards implementation related to three clinical KPI achievements categories: low, less than 60, moderate, 60 to 70, and higher performers, above 70.

It shows that the best-performing centers as per KPI achievement demonstrated better achievement in implementing the PCMH system as per NCQA assessment. Figure 1b shows that centers with longer adherence to the NCQA PCMH standards (average of 2018, 2021, and 2022) were enabled to better meet the clinical KPIs. Centers that achieved better adherence in 2022 to NCQA PCMH standards, compared with 2018 and 2021, achieved less in clinical KPI than those that were better in the 3 years. Figure 1a shows that the score of the NCQA PCMH assessment before the COVID-19 pandemic was better than that of 2021, indicating the major negative impact of the pandemic on quality of care, which was followed by an improvement in the recovery year of 2022 with centers showing high implementation of NCQA PCMH and CDC standards. Nevertheless, in terms of the 2022 clinical KPI, all the better-performing centers were superior to others in adherence to these standards.

**Figure 1b.**
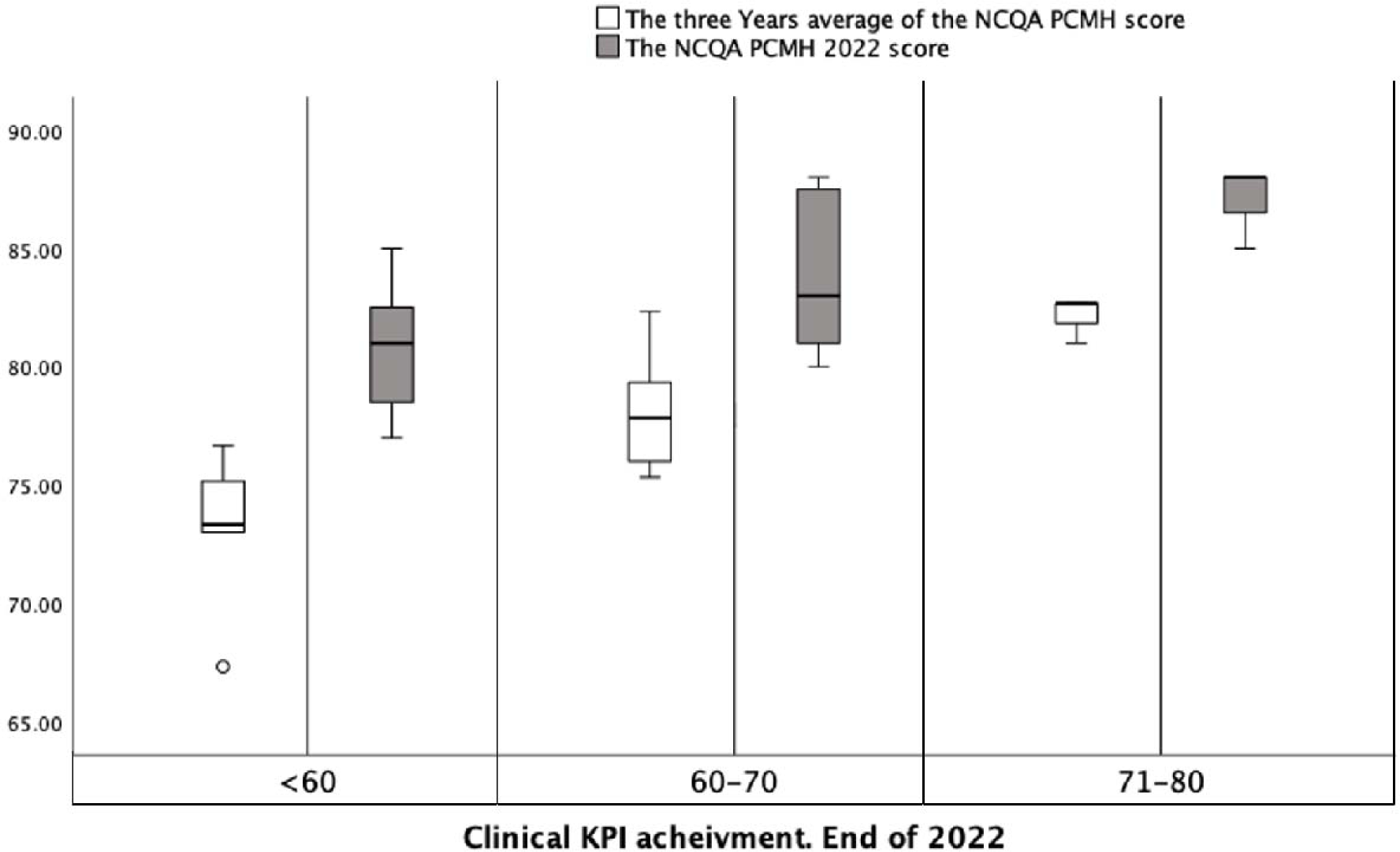
Centers’ adherence to the NCQA PCMH standards (average of the three years 2018, 2021, and 2022 NCQA PCMH scores) compared to 2022 NCQA PCMH performance.

Linear regression showed that better implementation of the NCQA PCMH and CDC programs was associated with better KPI outcomes and not related to changes in centers’ financial revenue. In the 2023 mid-year clinical KPI, care was superior in centers with better NCQA PCMH scores at the end of 2022, except in rural centers (B = 0.479 p = 0.044). However, rural centers demonstrated a similar association between the implementation of PCMH and clinical KPI achievement in the years before (Figure 2).

**Figure 2.**
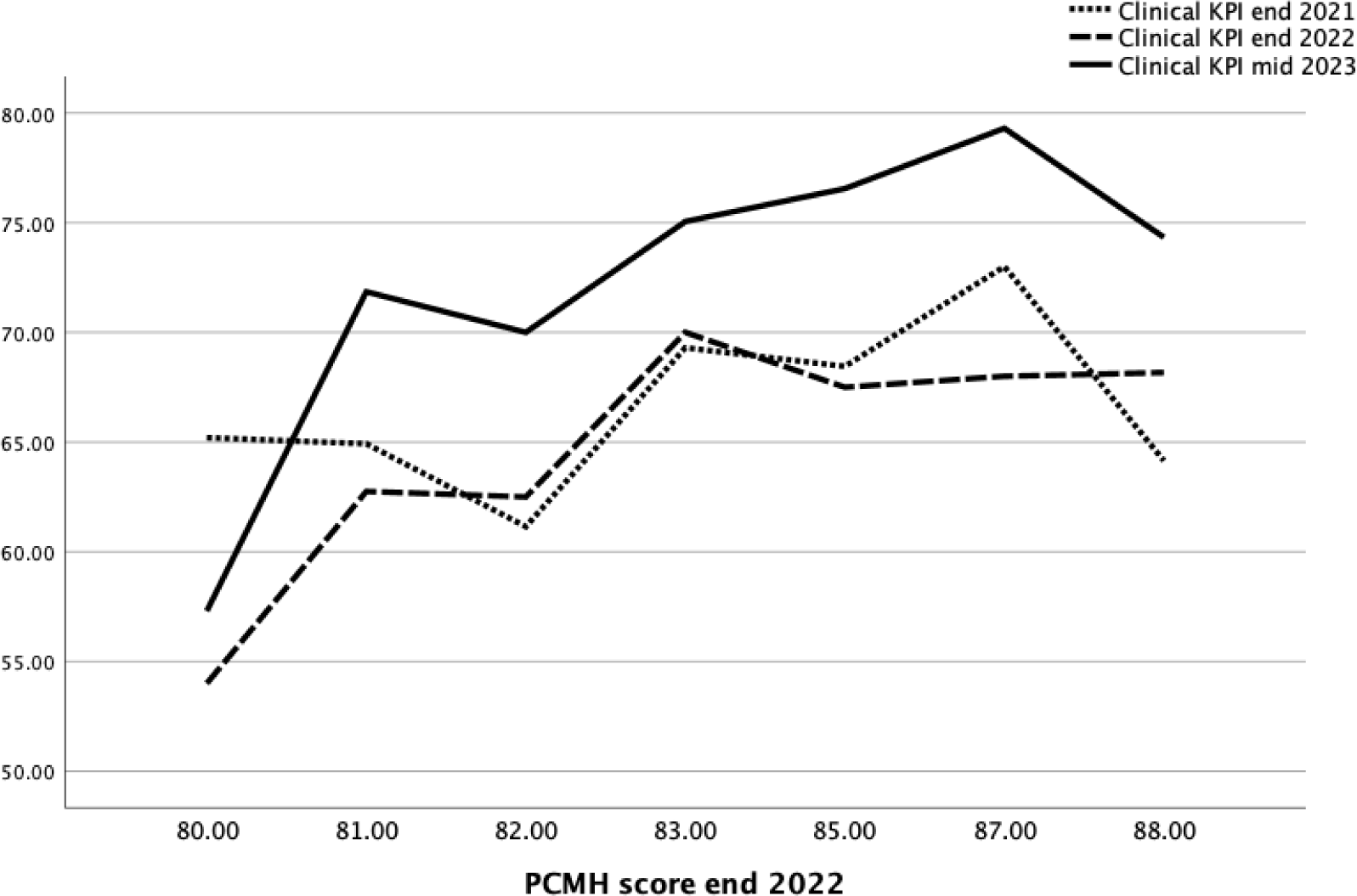
Clinical KPI average of all centers in relation to NCQA PCMH scores at the end of 2021, end of 2022, and mid-2023.

A significant association (B = 0.447, p = 0.03) was observed between the end of 2022 clinical KPI performance of all centers and the PCMH NCQA end-of-year 2022 score using linear regression. However, centers’ financial revenue, reflecting the financial implication, was not significantly associated with a better end-of-year clinical KPI (B = .209, p = 0.29). Similarly, through univariate linear regression, a higher CDC program assessment at the end of 2022 was significantly associated with centers that better performed in clinical KPI (B = 0.480, p = 0.013). The Chronic Diseases Program assessment at the end of 2022 was positively and significantly associated with the higher performance of centers regarding the NCQA PCMH standards (B = 0.647, p < 0.001).

### Correlation analysis of PCMH and CDC scores and outcome assessed (KPI and utilization)

The end of 2022 metrics (Table 2) analyzed by PCA emphasized the association between better outcomes in clinical KPI and adherence to NCQA PCMH and CDC standards. Significant correlations were observed between clinical KPI achievement in 2022 and the PCMH NCQA 2022 score (p = 0.004), preventive KPI achievement in 2022 (p = 0.028), higher centers’ financial revenue (p = 0.038), and higher CDC program score in 2022 (p = 0.013). The PCMH NCQA 2022 score significantly correlated with a higher centers’ financial revenue (p = 0.022) and a higher CDC program score in 2022 (p < 0.001). In addition, the CDC program score in 2022 significantly correlated with higher utilization, centers’ financial revenue (p = 0.008), more lab orders (p = 0.017), and more prescriptions (p = 0.004). Preventive services significantly correlated with centers with better clinical KPI but not any other variable.

**Table 2.**
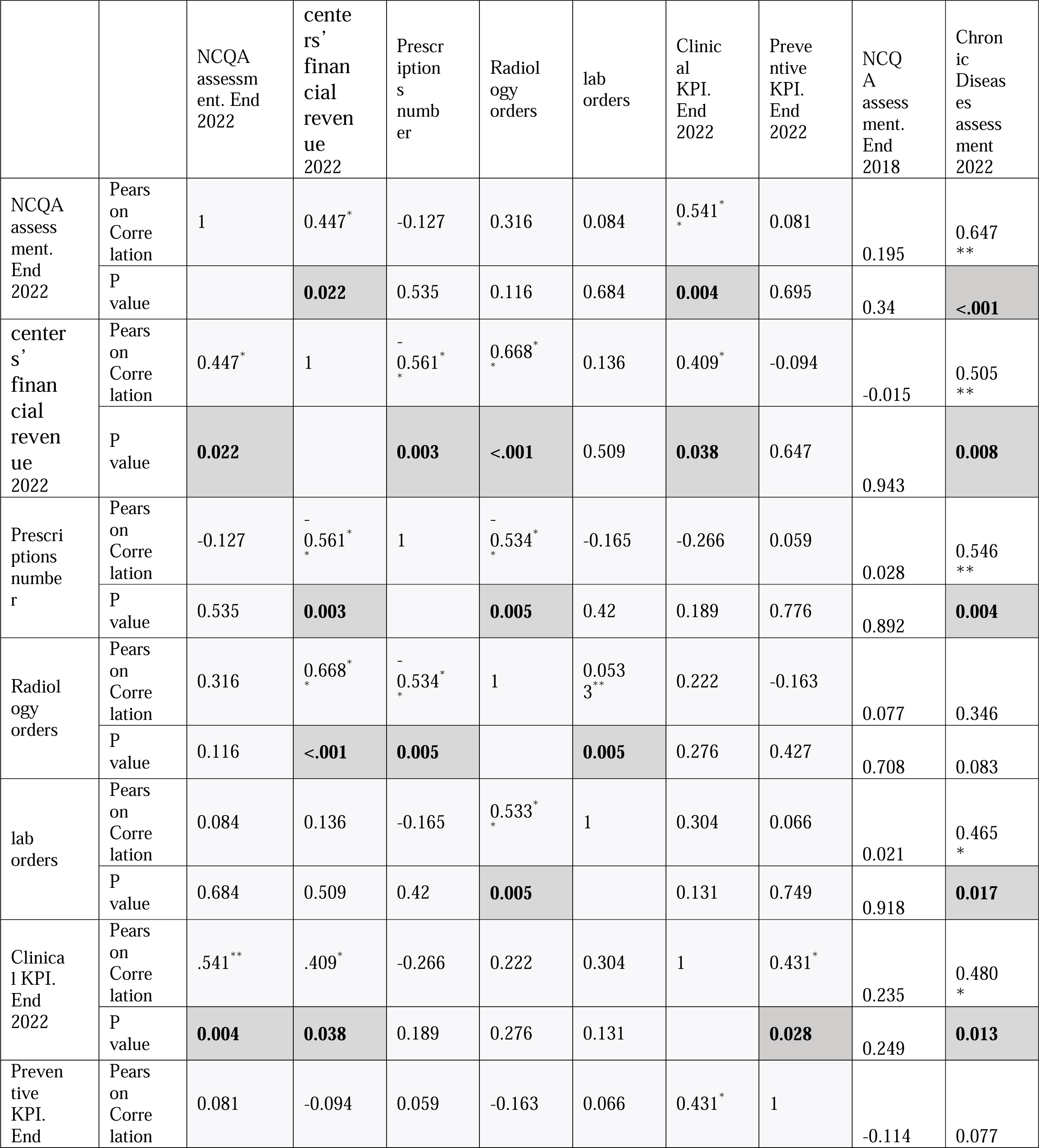
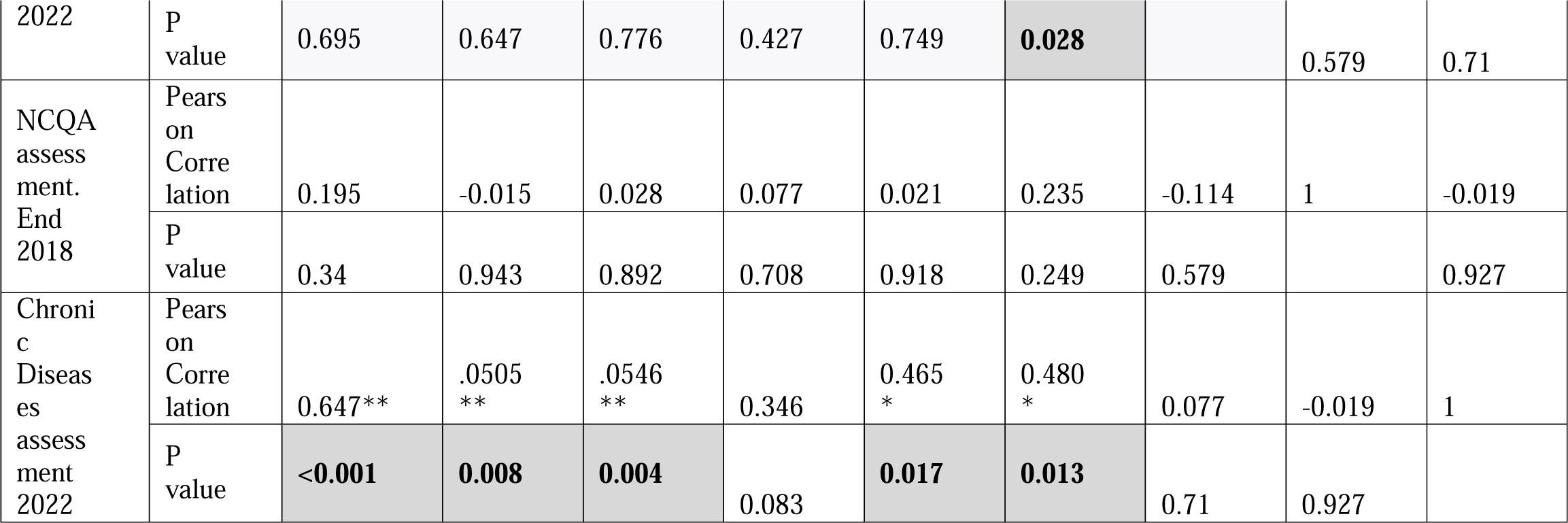
Correlations between the PCMH NCQA scores of the three years assessed, the volume of the services represented by the centers’ financial revenue, prescriptions, and radiological and laboratory requests, and the outcome KPI represented in the clinical and preventive KPIs in addition to chronic diseases care program assessment.

## Discussion

This paper reports 10 years of the real-world implementation of evidence-based standards, the NCQA PCMH, in an ambulatory care setting and over 18 years of a CDC management program in Abu Dhabi (Baynouna Al Ketbi et al., 2018; Baynouna et al., 2010). The positive association between PCMH recognition and clinical performance in healthcare centers was well established in quality of care, QoL, utilization, cost, and patient experience (NCQA, 2019) (John et al., 2020) (Glasgow, Whitesides, Nelson, & King, 2005). However, this study supports the effectiveness of its implementation in Abu Dhabi and reports sustainability over nearly 10 years with improvements in PCMH even in the less-performing centers. The clear effect of the COVID-19 pandemic was a decrease in adherence and care. Nevertheless, the superior care in centers with better implementation and adherence to standards during the pandemic, as per the repeated assessments, indicates significant effectiveness and the value of the structure and processes implemented.

Regression analysis showed that utilization, defined by patient volume, investigations, and prescription numbers, was not affected by the implementation of these programs. PCA revealed a positive influence on possibly an appropriate. Patients were recalled for the population health program, prediabetes, early hypertension detection, CKD, dyslipidemia, and stroke if they did not have a scheduled appointment. In addition, to the better implementation of the CDC program, this increase reflects appropriate, more scheduled, evidence-based care. There are many elements of the programs that require proactive planned care, such as panel management and coordination of population health programs. Part of it is to order investigations and adjust medications to close care gaps. Thus, an increase in centers’ financial revenue, investigations, and prescriptions could meet the clinical practice guidelines implemented in the disease, preventive, management, and population health programs.

When it comes to sustainability and progress in PCMH implementation, recognition and maintenance of that recognition is standard practice in the United States. However, in the current context, the commitment made by management and teams in the healthcare centers has been longstanding. Therefore, comparing this sustainability to others is challenging, but it remains a crucial area for study, as the benefits and return on investment could have played a role in fostering such commitment.

The pandemic significantly disrupted care delivery. Here, adherence to practice improvement strategies was affected in the centers. Notably, centers with better implementation of standards achieved better care outcomes during this period. Other countries reported the same impact of the pandemic on care delivery. As an example of corrective action, the Veterans Health Administration (VHA) launched the Preventive Health Inventory (PHI) program, a multicomponent care management intervention, including a clinical dashboard and templated electronic health record notes, to support primary care in delivering CDC and preventive care that the pandemic had delayed. A higher use of this multicomponent care management intervention was associated with improved quality-of-care metrics (Wheat et al., 2023). Therefore, the AHS, alongside the PCMH and CDC programs, provided support that was evident post-pandemic, as centers achieved better outcomes through the use of practice improvement initiatives to aid recovery.

The experience has unique characteristics. First, it describes a mixed intervention implementation with individual elements of the standards whose uptake is influenced by the distinctiveness of the centers and their team efforts. The level of PCMH implementation seemed to be a determining factor of the team efforts, as concluded by(Quigley et al., 2021). Advanced PCMH practices emphasized changes in the continuity of care, highlighting a focus on personal relationships rather than systemic change. Here, all the centers were on a shared journey as a network; however, analyzing the variability of efforts and their determinants could serve as a valuable research interest.

Second, while the PCMH standards are comprehensive and encompass numerous family medicine principles, the implementation of CDC programs in AHS since 2004 has been beneficial. The CDC program has been more prescriptive in structuring the flow of patients’ and teams’ responsibilities. Through the EMR, which serves as the foundation for population health programs, it connects to a vast pool of the population from which patients with chronic diseases are continuously identified. This could help improve adherence, as established and agreed-upon pathways and workflows can enhance the uptake of program strategies.

Rural centers did not differ from other centers in terms of the association between high PCMH scores and clinical KPI achievements. However, in 2023, there was an indication of a difference, with no association observed. More data regarding PCMH implementation in 2023 in rural centers can explain this finding. Gale et al. stated that although the readiness of the rural center for PCMH implementation varied, rural centers performed best on some NCQA PCMH standards, but many were challenging (Gale, Croll, & Hartley, 2015). Similarly, in Abu Dhabi, there was no difference in resources and infrastructure. However, owing to the low populations in certain regions, the teams were smaller, and care coordinators were not available in certain centers and were shared in others. Furthermore, the population demographics differed, and this affected the clinical KPI performance of the rural centers.

This experience reporting has a limitation that is similar to similar experiences in healthcare services research: the difficulty of attributing effectiveness to a specific implemented element. Which were the most impactful elements? This was a challenge in studies in the US, where no PCMH implementation resembled others. Bodenheimer (2022) noted that “because PCMH is a diffuse collection of initiatives rather than a focused intervention, evaluation is difficult” (Bodenheimer, 2022). Flieger (2017) stated, “If you have seen one medical home, you have seen one medical home” (Flieger, 2017). Research in this area could be more easily conducted in the financial domain, as posited by Burton et al. (2020), who identified components of the PCMH model of care associated with lower spending and utilization among Medicare beneficiaries (Burton et al., 2020). However, identifying the same for the quality of care and patient experiences may not be easy as they are not as easy to quantify. This is more complex in unique settings, such as Abu Dhabi, as the confounders differ, and it is the only one in the region; therefore, generalizability is a risk. Nevertheless, the transferability of the core elements of the standards and the success of implementation, sustainability over time, and positive association with key clinical outcomes encourage the better delivery of family medicine services.

The improvement in patient care associated with better adherent centers has reached international benchmarking levels in certain recognition programs, such as diabetes, stroke, and heart disease (NCQA, 2023b) (NCQA, 2023a). However, inertia was observed, probably attributed to factors, which can only be confirmed by studies. The open-access system is allowing patients to break their continuity with their PCP, and care is interrupted by many options from different providers, which interfere with the care planned in CDC clinics. This open-access system facilitates timely accessible care; however, that is to be weighed against the duplication of services and less adherence to appointments and PCP continuity, with patients visiting urgent-care clinics for routine chronic care. The lack of evidence on reducing visits in this study may be linked to the fee-for-service system. No association was observed between the PCMH score and financial outcome even when controlling for rural centers. This variability needs to be studied for determinants of health-seeking behavior and physician practice. In the US, where the fee for services is highly implemented, there is migration out of the system. This is because of the high-cost, low-value outcomes and more interest in initiatives in primary care, particularly PCMH, which showed better savings in centers implementing PCMH (Sinaiko et al., 2017). Friedberg et al. (2018) observed that practicing physicians find it difficult to keep up with the proliferation of models, and payment models have become increasingly complex since 2014, with practices investing in understanding these complex alternative payment systems. Furthermore, they discovered that risk aversion was prominent among physician practices (Friedberg, Peggy G. Chen, Molly M. Simmons, Tisamarie B. Sherry, Peter Mendel, Laura Raaen, Jamie Ryan, Patrick Orr, Carol Vargo, Lindsey Carlasare, Christopher Botts, and Kathleen Blake, 2018). Neumann (2019) used Medical Expenditure Panel Survey data to analyze the relationship between primary care spending, PCMH implementation score, and health outcomes in 29 states and discovered that greater primary care investment led to better outcomes. Furthermore, PCMHs can help to reduce costs and improve population health. (Neumann, 2019)

The challenges of fulfilling the core functions of primary care, first contact, continuity, coordination, and comprehensiveness are within the confines of the fee-for-service (K. John McConnell, 2019). However, with the present PCMH-based structure and processes and CDC team in all centers, as well as informational continuity and care coordinators completing tasks, the center teams maintained continuity to achieve results.

This study showed the value of investments in PCMH standards with a focus on structured care in CCM-based disease management programs for CDC related to a better quality of care and utilization. In addition to a possible higher interest in change toward better healthcare delivery post-pandemic, a targeted assessment is required.

The strength of this reported experience is sustainability despite the lack of recognition similar to the US system. A US study concluded that small and medium-sized practices may experience difficulty with the financial burden of PCMH recognition, and transformation is disruptive to practices, requiring the commitment of leadership and personnel. However, their value requires policies that recognize and meet the requirements of on-site practice leaders to promote primary care practice transformation (Lieberthal, Payton, Sarfaty, & Valko, 2017) (Goldman et al., 2018) (Qureshi, Quigley, & Hays, 2020). Thus, any transformation toward PCMH is recommended, although the value of certification and recognition cannot be determined from this study. In AHS, the sustainable leadership decision to support its implementation shows the commitment of the healthcare team to adhere to standards, even during the pandemic. Importantly, the family medicine-based care (Starfield, Shi, & Macinko, 2005) can never be achieved without the PCP. The link to the patients in their panel is the center of all elements in the programs described. In AHS, the empanelment is facilitated through the EMR, and reports and care coordination were easier with the PCP identified for each patient and each PCP having their panels.

A unique sustainability attribute in this country’s context is that practice improvement is centralized and governed in coordination with key departments to facilitate and control its quality and implementation. Davy et al. supported this, emphasizing the commitment of leaders and spreading awareness (Davy et al., 2015). Finally, this study highlights the clear processes and outcomes followed over time. In many studies, sustainability has no explicit definitions of outcome variables; thus, research cannot accumulate or disconfirm findings on sustainability predictors (Scheirer & Dearing, 2011).

## Conclusion

The Abu Dhabi AHS investment and success in implementing evidence-based interventions demonstrated good-to-excellent implementation of both programs. Excellent sustainability was evident over the years, despite the noticeable decrease in PCMH scores and KPIs during the COVID-19 pandemic, which was followed by the highest score since the program began. The setting and strategies discussed underscore significant issues for sustainability research and provide evidence of the importance of maintaining interventions over time.

## Declarations

### Ethical approval and consent to participate

The study was approved by the Department of Health-Abu Dhabi IRB. All methods were carried out under relevant guidelines and regulations.

### Consent statement in the Ethics approval and consent to participate

Informed consent was waived by the Department of Health-Abu Dhabi IRB as the study was designed for retrospective data gathered as part of patient care and organization quality program and anonymized at analysis.

### Consent to publish

Not Applicable.

### Availability of data and materials

Data is available on request sent to the corresponding author.

### Author contributions

LB conceptualized, analyzed data, and wrote the manuscript. NN assisted in statistical analysis and reviewed the manuscript.

### Conflicts of Interest and Source of Funding

All authors declare no conflicts of interest, and no funding was received for this study.

## Data Availability

Data is available on request sent to the corresponding author.

## Notes

### Competing Interest Statement

The authors have declared no competing interest.

### Funding Statement

none

