## Appendix 1 for "Sustainability of Evidence-Based Practice Improvement Programs in Abu Dhabi Ambulatory Healthcare Services for more than a decade and During the COVID-19 Pandemic"

Appendix (1) Population Health Programs mandated by the government and the AHS introduced Chronic Diseases and PCMH program elements.

| **Population based Program Government** | **Target** |
| --- | --- |
| Adult cardiovascular screening program | Adults |
| Adult Comprehensive preventive screening | Adults |
| Well Child Care | Children less than 2 years |
| Premarital | adults |
| University Screening | adults |
| Drivers license fitness | adults |
| Occupational fitness | adults |
| Visa renewal screening | adults |
| Travel medicine | Travelers |
| Antenatal care | Pregnant women |
| School screening | School students |
| **Chronic Diseases Care CDC management program** |  |
| Chronic diseases Care | All population with chronic illness |
| Diabetes Recognition Program | Adults |
| Osteoporosis Program | Adults |
| Asthma Recognition Program | Children and adults |
| Hypertension Recognition Program | Adults |
| Mental health Recognition Program (Depression, Anxiety, Autism, Dementia) | Children and adults |
| Chronic Kidney Disease CKD Recognition Program | Adults |
| Pre-Diabetes Program | Adults |
| Hypertension detection program | Adults |
| Complex patients |  |
| Dyslipidemia Program |  |
| **Population stratification** |  |
| By disease (Problem list and sources above) | Children and Adult |
| By AAFP modified Risk stratification levels: (judged by Physician) | Children and Adult |
| By PCP panel: (Choice of the patient) | Children and Adult |
