## Appendix 2A for "Sustainability of Evidence-Based Practice Improvement Programs in Abu Dhabi Ambulatory Healthcare Services for more than a decade and During the COVID-19 Pandemic"

Appendix 2-A (new) Summary of PCMH 2017 standards with core and electives points and examples of evidence to meet the standards elements as implemented over the years in AHS

| 2017 PCMH Self-Assessment | | |  |
| --- | --- | --- | --- |
| **Concepts, Competencies, and Criteria** | **Self Assessment Columns** | | **Examples of evidence of implementation of the standards Elements** |
|  | **CORE Need all 40 core points** | **ELECTIVE Need 25 of 60 credits from 5 of the 6 Concepts** |  |
| I. **Team-Based Care and Practice Organization (TC)** | **5 core** | **7 credits** |  |
| Competency A: The practice is committed to transforming the practice into a sustainable medical home. Members of the care team serve specific roles as defined by the practice’s organizational structure and are equipped with the knowledge and training necessary to perform those functions. | 2 core | 5 credits | 1. Regular Baytona AlTebi Meetings and Annual Baytona AlTebi Collaborative 2. Hierarchy within the team 3. Team meetings and Huddle record 4. Order by Protocol 5. Mental Health Distinction Program agenda 6. Baytona Al Tebi letter and leaflets. Marketing material of AHS |
| Competency B: Communication among staff is organized to ensure that patient care is coordinated, safe and effective. | 2 core | 2 credits |  |
| Competency C: The practice communicates and engages patients on expectations and their role in the medical home model of care. | 1 core | No elective credits |  |
| **II. Knowing and Managing Your Patients (KM)** | **10 core** | **22 credits** |  |
| Competency A: Practice routinely collects comprehensive data on patients to understand the background and health risks of patients. Practice uses information on the population to implement needed interventions, tools and supports for the practice as a whole and for specific individuals. | 3 core | 6 credits | 1. Comprehensive Health Assessment tools 2. Home monitoring page to be completed by CDC nurse 3. Conducts depression screenings for adults and adolescents using a standardized tool. 4. Dental department agenda 5. Social Determinants of Health 6. Health Literacy Tools and interventions 7. Panel Management 8. Implemented recognition programs 9. Medication safety and adherence 10. CKD alert 11. Evidence-Based Decision support 12. Community resources use and outreach collaboration   Collaborations with other centers and hospitals in Knowledge transfer and case conferences |
| Competency B: The practice seeks to meet the needs of a diverse patient population by understanding the population’s unique characteristics and language needs. The practice uses this information to ensure linguistic and other patient needs are met. | 2 core | 1 credits |  |
| Competency C: The practice proactively addresses the care needs of the patient population to ensure needs are met. | 1 core | 2 credits |  |
| Competency D: The practice addresses medication safety and adherence by providing information to the patient and establishing processes for medication documentation, reconciliation and assessment of barriers. | 2 core | 5 credits |  |
| Competency E: The practice incorporates evidence- based clinical decision support across a variety of conditions to ensure effective and efficient care is provided to patients. | 1 core | No elective credits |  |
| Competency F: The practice identifies/ considers and establishes connections to community resources to collaborate and direct patients to needed support. | 1 core | 8 credits |  |
| **III. Patient-Centered Access and Continuity (AC)** | **7 core** | **8 credits** |  |
| Competency A: The practice seeks to enhance access by providing appointments and clinical advice based on patients’ needs. | 5 core | 4 credits | 1. Appointment system 2. Patients Portal 3. PCP panel size , adjustment and change 4. School Nurses Panel 5. PCP continuty monitoring 6. Panel Coordinator   Panel szie review and reconciliation |
| Competency B: Practices support continuity through empanelment and systematic access to the patient’s medical record. | 2 core | 4 credits |  |
| **V. Care Management and Support (CM)** | **4 core** | **6 credits** |  |
| Competency A: The practice systematically identifies patients who may benefit from care management. | 2 core | 2 credits | 1. Workflow in structured care (CDC, mental health and preventive, ANC, Women’s Health) 2. Triaging ( Triage nurse ) 3. Population health early recognition programs 4. Risk stratification 5. Care plan documentation, guidelines and audits   Self Management |
| Competency B: For patients identified for care management, the practice consistently uses patient information and collaborates with patients/families/ caregivers to develop a care plan that addresses barriers and incorporates patient preferences and lifestyle goals documented in the patient’s chart. | 2 core | 4 credits |  |
| **VI. Care Coordination and Care Transitions (CC)** | **5 core** | **24 credits** |  |
| Competency A: The practice effectively tracks and manages laboratory and imaging tests important for patient care and informs patients of the result. | 1 core | 3 credits | 1. Investigation tracking by centers and central monitoring KPI 2. Safety KPI in Investigation tracking 3. Tele for orderset defaulter and for results 4. Orderset and decision support for lab and imaging 5. Referal tracking and central monitoring 6. Referal criteria guide 7. Physcians directory 8. Physcians Performance information 9. Referals Providers letters 10. Referrals Feedback 11. School Nurses E-referral 12. Message center notification: ED and discharge alerts and its workflow and audits 13. Transition in or out of practice (peds, between centers, to home care)   Home care: Transition of care |
| Competency B: The practice provides important information in referrals to specialists and tracks referrals until the report is received. | 1 core | 14 credits |  |
| Competency C: The practice connects with health care facilities to support patient safety throughout care transitions. The practice receives and shares necessary patient treatment information to coordinate comprehensive patient care. | 3 core | 7 credits |  |
| **VII. Performance Measurement and Quality Improvement (QI)** | **9 core** | **16 credits** |  |
| Competency A: The practice measures to understand current performance and to identify opportunities for improvement. | 4 core | 4 credits | 1. Quality Reports review and development 2. Depart summary 3. CAHPS, PACIC 4. KPI review   KPI dashboard |
| Competency B: The practice evaluates its performance against goals or benchmarks and uses the results to prioritize and implement improvement strategies. | 4 core | 5 credits |  |
| Competency C: The practice is accountable for performance. The practice shares performance data with the practice, patients and/or publicly for the measures and patient populations identified in the previous section. | 1 core | 7 credits |  |
