## Appendix 2B for "Sustainability of Evidence-Based Practice Improvement Programs in Abu Dhabi Ambulatory Healthcare Services for more than a decade and During the COVID-19 Pandemic"

Appendix 2 B Chronic Diseases Care Program Audit Components

| No. | 1.Empanelment | Score (0,1,2) | Evidence |
| --- | --- | --- | --- |
| 1.1 | Schedule implemented ( covered by one doctor every day) | 2 | send schedule with CDC assignment) |
| 3.1 | Chronic Diseases Clinic is scheduled every day | 2 | Salamatak daily appointment, Report monitoring CDC slot% |
| 4.1 | seen by PCP ( Continuity in PCP) | 2 | Example 10 |
| 5.1 | Use PCP to allocate new patients to PCP | 2 | 1.Empanelment report |
|  |  |  | BA reporrt |
| 6.1 | Appointment given according to PCP | 2 | EMR Check |
| 7.1 | Clean up method used (Number change BA rreport)tracking sheet | 2 | at visit (sheet) |
| 8.1 | No show monitor | 2 | (referral tracking) KPI (excel), month of reschedule Q1, Q2) |
| 9.1 | Panel review by manager with action plan | 2 | Panel Size adjustment: Action plan afterr BA email. |
|  | 2.Teams based care |  |  |
| 1.2 | Teamlet identified in for Complex care population. (for complex patients care) | 2 | CME attendance |
| 1.2 | CDC Nurse schedule , CDC Nurse backup | 2 | CME attendance |
| 2.2 | CDC-training attended | 2 | Case management and BA monthly webinar (attendance match) |
| 3.2 | Huddle | 2 | (Huddle time fixed in schedule, daily CDC and Weekly CDC-AT) |
| 4.2 | Pre-Planned forms used/ Communication slip | 2 |  |
| 5.2 | Evidence of teamlet training for the population identified | 2 | List |
| 6.2 | Teamlet EMR competency used and staff trained. | 2 | Super user training staff in centre |
| 7.2 | Health Educator assigned /self-management Nurse | 2 | Assigned Nurse (name) |
|  | 3.Population Management |  |  |
| 1.3 | Health Maintenance random audit result (Panel Management) | 2 | EMR audit |
| 2.3 | Documentation in the right comprehensive assessment form. | 2 | EMR audit |
| 4.3 | Dementia Screening | 2 | BA report/  EMR audit |
| 4.3 | Depression and anxiety Screening | 2 | BA report/  EMR audit |
| 6.3 | Smoking screening from 13 years evidence cases and have Smoking Clinic | 2 | BA report/  EMR audit |
| 7.3 | Know about the Population Health program | 2 | BA report |
| 13 | Recall all those in PH list | 2 | Lists sent |
| 13 | How many see of all those in PH list | 2 | Lists sent |
| 8.3 | Asthma assessment done (evidence ) | 2 | Recall from PCP Panel to CDC (telecoslt/F2F). |
| 9.3 | Complex patient identified and followed | 2 | Complex patients List sent |
| 10 | Panel management to review gaps in care. | 2 | Audit on CC |
| 11 | Order set use | 2 | Report |
| 12 | Know about the Use the medical calculators | 2 | Audit |
| 13 | Use of specified Population page (ANC ,WCC,….) pregnancy summary | 2 | EMR audit |
| 13 | School Screening students given appointment to PCP in clinic | 2 | Tele-Consult for Peds Panel Mx |
| 13 | Identified obese children and ordered the metabolic panel &referral to Dietitian | 2 | Lists sent |
|  | 4.Care management and support |  |  |
| 1.4 | Medication reconciliation above 90% | 2 | Report |
| 2.4 | Self-management | 2 | Audit Pharmacy disspense of Devices (BP, CGM, PEF) |
| 3.4 | Assess medication understanding | 2 | Pharmacy tools (method) |
| 4.4 | Health literacy assesment tool used | 2 | Brown bag/HL forms GF |
| 6.4 | Assess patient response to treatment between visits | 2 | of calling |
| 7.4 | New medication counseling | 2 | Pharmacy tools (method) |
| 8.4 | PACIC or CAHPS done for more than 50 patient in the last quarter do more (10/month ) | 2 | Action plan for Patients experince |
| 9.4 | AHS center counsel | 2 | Documented meetings annd action plan |
|  | 5.Care coordination |  |  |
| 1.5 | Use investigation tracking report | 2 | Report utilization |
| 2.5 | Use message pool | 2 | Discharge, ED visits |
| 3.5 | Use message center by doctors | 2 | Audit on doctors center by manager/CL |
| 4.5 | follow up of positive labs documented in excel | 1 |  |
| 5.5 | Use referral tracking report | 2 | Excel review |
| 6.5 | Follow on messages of admissions | 2 | Discharge, ED visits |
| 7.5 | Depart Summary use | 2 | Report |
| 8.5 | Provider Letter use | 2 | Report |
|  | 6. Operational |  |  |
| 2.6 | Daily structured clinic (CDC) | 2 | EMR Check |
| 3.6 | Clinic objectives available as CDC | 2 |  |
| 4.6 | Doctors see all populations in the structured clinics/ | 2 | EMR audit |
| 5.6 | Asthma daily clinic appointment /Schedule | 2 | EMR audit |
| 6.6 | One week fixed schedule for Drs/Schedule for the year | 2 | schedule to be sent |
| 7.6 | Academic slot utlised : fixed for each residents if applicable | 1 | EMR audit |
| 8.6 | Mental health positive cases managed and documented | 2 | EMR audit |
| 9.6 | Referral Patients to Dietitian & Health Educator | 2 | EMR audit |
| 12 | Internal referral feedback by specialist | 2 | Report |
|  | 7.PCP Audit |  |  |
| 1.7 | Fixed weekly Schedule for all PCP | 2 | schedul/EMR audit |
| 2.7 | Work on clean up from panel clean-up report. | 2 | Report |
|  | 8.Teleconsultation Audit |  |  |
| 1.8 | Panel clean-up through the teleconsultation | 2 | excel review |
| 2.8 | Fixed weekly Schedule for all PCP | 2 | schedul/EMR audit |
| 3.8 | For auditing PCP tele-visit. (Follow up with the same PCP | 2 | EMR audit |
| 8.Teleconsultation Audit | |  |  |
|  | 9.Portal Utilization Audit |  |  |
| 1.9 | Education for staff | 2 |  |
| 2.9 | Education for patient how to use | 2 |  |
| 9.Portal Utilization. Audit | |  |  |
|  | 10.Patient Education |  |  |
| 1.1 | Health literacy Health education | 2 | know about |
| 2.1 | DOH /SEHA  Campaign - Educating the patients about Diseases | 2 |  |
| 3.1 | Self-management Nurse between visit follow-up | 2 | know about |
| 10.Patient Education | |  |  |
| Total scores: | | 116 |  |
| Percentage out of 100 | | 100 |  |
