## Appendix 3 for "Sustainability of Evidence-Based Practice Improvement Programs in Abu Dhabi Ambulatory Healthcare Services for more than a decade and During the COVID-19 Pandemic"

**Appendix 3: Summary of processes, clinical, and preventive Key Performance Indicators monitored on a monthly basis.**

| **POPULATION HEALTH AND DISEASE MANAGMETN RECOGNISTION PROGRAMS** | **TARGET** |
| --- | --- |
| CKD Recognition Program: % of NSAIDs used in CKD2 within the last 6 months , % of NSAIDs use in CKD34 in the last 6 months , % of patient with Proteinur ia and on ACEIorA RB , % CKD2 patients with hyperlipiemia and on Statin, % CKD34 patients with hyperlipiemia and on Statin, % CKD34 patients with SBP>140, % CKD34 patietns with DBP>90 | **75%** |
| DM Recognition Program: percentage of diabetic patients with: HbA 1C Poor Control >9.0%, HbA 1C Control <8.0%, HbA 1C Control <7.0%, Blood Pressure Control ≥140/90 mm Hg, Blood Pressure Control <140/90 mm Hg, eye examination, Smoking cessation referrals, LDL control >=130, LDL control >=100, foot examination. | **75%** |
| Heart / Stroke Recognition Program: Blood Pressure Control <140/90 mmHg, Smoking Status /Cessati on advice or treatment, complete Lipid Profile, LDL Control <100 mg/dl , Use of aspirin or another anti-thrombotic Statin use. | **75%** |
| Asthma patients who had their Influnza vaccination within a year | **75%** |
| Hypertension recognition Program: patietns with hypertension with: BP less than 140/90, LDL done, nephropathy Assessment done | **75%** |
| Smoking Recognition Program: % of patients attending the center who had smoking status assessed, % of smokers referred to smoking cessation clinics | **75%** |
| Obesity ADULT : order set within EMR that includes recommended labs and referrals which was ordered for patients with obesity annually | **75%** |
| Obesity CHILD: order set within EMR that includes recommended labs and referrals which was ordered for patients with obesity annually | **75%** |
| Mental health Screening (Depression): PHQ9 done for adults annually | **75%** |
| Mental health Screening Program (Anxiety) GAD7 done for adults annually | **75%** |
| **PCMH Operational KPI** |  |
| CDC Visit Type (DM & HTN) | **90%** |
| Percentage of Diabetic CDC Follow Up Appointments given to all chronic diseases patients | **98%** |
| Percentage of Diabitec Dental Referral | **50%** |
| CDC consultation delivered through Telemedicine | **25%** |
| CDC Telemedicine Done by the patient PCP | **50%** |
| Percentage of patients with Primary Care Physician (PCP) assigned | **98%** |
| Percentage of patients with Primary Care Dentist (PCD) assigned | **90%** |
| Percentage of Medication Reconciliation done in all consultations | **90%** |
| Percentage of Medication Reconciliation done in CDC consultation | **98%** |
| Referral: Percentage of provider letter Initiation done | **80%** |
| Referral Feedback received by the refering physician | **50%** |
| Percentage of Depart Summary Generated | **50%** |
| Care Coordination and Care Transition: Investigation tracking report utlisation by the care coordinator | **100%** |
| Percentage of the following investigations’ results that were endorded by the ordering physcian:  Follow Up Of Screening Test Result (Mammogram)  Follow Up Of Screening Test Result (Pap Smear)  Follow Up Of Screening Test Result (Dexa Scan)  Follow Up Of Screening Test Result (Colon Screening) | **100%** |
| Percentage of Risk Stratification done | **90%** |
| **Population Health Recall** |  |
| Pre Diabetic Recall List | **75%** |
| Hypertension Early Detection Recall List | **75%** |
| CKD Stage 2 Recall List | **75%** |
| Dyslipidemia Recall List | **75%** |
| Complex Recall List (Diabetic>9,Hypertentsion >140, Stroke all, CKD Stage 3-5) | **75%** |
| **PCMH Utlisation** |  |
| Call after admission or ER visit (Message Pool From Centers) | **75%** |
| PCP Panel Report 40 patient Tracking (Adult/Child) | **75%** |
| PCP Panel Size (Biannual) | **75%** |
| **PCMH Patient experience** |  |
| CAHPS % done (replaced by new organisation patient experince program) | **20 (10 adult, 10Peds)** |
| PACIC % done | **10%** |
| **Overall NCQA (interim in June, End of Year December)** |  |
| NCQA score Core |  |
| NCQA Elecetive |  |
